# Views about integrating smoking cessation treatment within psychological services for patients with common mental illness: a multi-perspective qualitative study

**DOI:** 10.1101/2020.02.18.20024596

**Authors:** Gemma Taylor, Katherine Sawyer, David Kessler, Marcus Munafò, Paul Aveyard, Alison Shaw

## Abstract

**Background:** Smoking rates are significantly higher in people with mental health problems, compared to those without. Negative attitudes towards smoking cessation are widespread in inpatient settings towards patients with severe and enduring mental illness. It is not clear if the same attitudes operate in psychological services towards people with common mental illness. We aimed to understand the concerns and views that patients, therapists, and smoking cessation practitioners may have about integrating smoking cessation treatment into psychological treatment for common mental illness and how these concerns may be overcome.

**Methods:** Thematic analysis of 23 in-depth interviews. Interviews took place in Improving Access to Psychological Therapies (IAPT) and smoking cessation services in England. Participants were 11 psychological wellbeing practitioners (PWPs), six IAPT patients with common mental illness, and six smoking cessation advisors.

**Outcomes:** None.

**Findings:** IAPT patients reported psychological benefits from smoking, but also described smoking as a form of therapeutic self-harm. PWPs seem positive towards smoking cessation treatment for people with common mental illness. IAPT PWPs and patients accept evidence that smoking tobacco may harm mental health, and quitting might benefit mental health. PWPs report expertise in helping people with common mental illness to make behavioural changes in the face of mood disturbances and poor motivation. IAPT appears to be a natural environment for smoking cessation intervention. PWPs felt confident to offer smoking cessation treatments to IAPT patients, but thought that a reduction in caseload was required to deliver smoking cessation support in an already pressed service.

## BACKGROUND

*“I was signed off work with depression. I was asked if I smoked and I was encouraged not to try and quit… I was given anti-depressants instead”*

> − Smoker with depression, aged 37

Smoking is the world’s leading cause of preventable illness and death^1^. One in every two smokers will die of a smoking-related disease, unless they quit^2,3^. Globally, smoking prevalence has decreased from 29% during the 1990s to about 15% in recent years^4^. However, smoking rates in people with common mental illness, like depression and anxiety, are at least twice the rate of those without common mental illness. For example, in the UK 34% of people with depression, and 29% of people with anxiety smoke^5^. This population is more heavily addicted, suffers from worse withdrawal^6^, and has a 19% reduction in the odds of achieving abstinence when making a quit attempt^7^, but are as motivated to do so as the general population^8^. These differences reduce life-expectancy in people with common mental illness when compared to the general population (mortality rate ratio, 1.92 (95% CI: 1.91 to 1.94)^9^.

One major barrier to implementing cessation treatments in this population is the widely held misconception that smoking improves mental health and that quitting may interfere with mental disorder treatment, and that smoking should be addressed once mental health has improved^10^. However, there is no clear reason why mental illness and tobacco addiction cannot be addressed simultaneously, and no evidence that stopping smoking causes psychological harm^11^. Conversely, there is growing evidence that smoking may worsen mental health through the tobacco withdrawal cycle^12^, and that stopping smoking may improve mental health, an effect size equal to anti-depressant treatment^11^.

A Cochrane review of smoking cessation interventions for people with current and historical depression found that adding psychosocial mood management to usual smoking cessation treatment (e.g., nicotine replacement therapy) increased smoking cessation rates in people with depression compared to usual smoking cessation treatment alone, risk ratio of 1.47 (95% CI: 1.13 to 1.92)^13^. Smoking cessation support could be appropriately placed in psychological services whereby people who want help to quit smoking would be offered the option to receive integrated psychological treatment for their mental health difficulties and tobacco addiction.

Sheals and colleagues conducted a systematic review, meta-analysis and qualitative synthesis of mental health professionals’ attitudes to treating smoking cessation in people with mental illness. They included 38 studies involving 16,369 mental health professionals^10^. The most commonly held attitudes amongst professionals were that patients were not interested in quitting, and that quitting smoking was too much for patients to take on. Results indicated a culture of smoking as ‘the norm’ and a perception that cigarettes were a useful tool for patients and staff. What is clear from Sheals’ et al’s review and other research is that negative attitudes towards smoking cessation are widespread in inpatient settings where professionals work with patients who have severe and enduring mental illness (i.e. psychosis), and in institutions that operate using predominately medically-based treatment models (i.e. hospitals)^10,14,15^. But what is not clear is whether or not these attitudes are widespread in institutions that work with patients with common mental illness, or in institutions that operate using predominately psychologically-based treatment models (i.e. psychological therapies services).

In England, people with common mental illness are usually referred to a community-based psychological therapies service, known as ‘Improving Access to Psychological Therapies (IAPT)’. In IAPT, patients receive cognitive behavioural therapies to improve mood symptoms and quality of life. IAPT could deliver smoking cessation treatment alongside CBT for common mental illness, but currently does not. Therefore, the aim of this qualitative study was to understand whether it was possible to integrate smoking cessation treatment into psychological therapies for people with common mental illness by understanding the relevant concerns of patients and staff.

### Research objectives

The specific objectives of the study were to:

1. Understand patient experiences of comorbid smoking and common mental illness (i.e., depression/anxiety);
2. Understand patient views about treatments for tobacco addiction and common mental illness, including parallel treatment of both;
3. Understand health professional’s knowledge and views of parallel treatment of tobacco addiction and common mental illness, including pharmacotherapy as an aid to smoking cessation.
4. Collect data to inform a potential smoking cessation intervention for integration into psychological therapies services (IAPT).

## METHODS

The protocol for this study has been registered on OSF (https://osf.io/z7vsy/), and we have followed COREQ reporting guidelines^16^. Ethics approval for this study was received from the NHS Research Ethics Committee on 13 July 2017 (Reference 17/WM/0251).

We conducted semi-structured interviews with IAPT psychological wellbeing practitioners (PWPs) and patients, and stop smoking service advisors.

### Sampling and recruitment

We recruited participants from IAPT services and smoking cessation services in England until we generated adequate information power, as defined by Malterud and colleagues^17^.

Participants were all aged ≥18-years. PWPs and smoking cessation advisors were recruited using a snowballing strategy at the local service level. We interviewed a range of males and females, including those were newly qualified (at least 1 year) or who were more experienced in their role (>2 years). IAPT patients were recruited by PWPs during IAPT appointments, using a purposive approach to ensure that participants had with a variety of common mental illness (all treatable in IAPT).

IAPT PWPs were non- or ex-smokers. Smoking cessation advisors had provided smoking cessation treatment to people with mental disorders, and were employed in a National Centre for Smoking Cessation Training (NCSCT) trained stop smoking service. IAPT patients had a current form of depression and/or anxiety, were currently receiving IAPT treatment or had completed treatment within a year of the interview, and had smoked daily for at least a year.

### Data collection

Interviews were conducted between September 2017 and April 2018. Participants were interviewed in-person or by telephone. All interviews were audio recorded and lasted typically 60 minutes.

Topic guides were used to assist questioning during semi-structured individual interviews (Appendix) with flexibility to reflect emergent findings. The interviewer (GT) used open-ended questioning to elicit participants’ own experiences and views and participants were asked to provide examples to avoid reliance on ‘hypothetical’ accounts.

Data were transcribed by a third-party service. To ensure quality of data transcription a researcher did a 50% check of audio data against the transcripts.

Participants were not paid for their contribution to the study, but were provided with sustenance during the interview, and travel costs were reimbursed.

### Data analysis

GT and KS led the analysis with support from AS. The data were analysed using a thematic approach, following guidance outlined by Braun and Clarke^18^; thematic analysis allows for both anticipated themes (i.e., deductive coding) and emergent themes (i.e., inductive coding).

We used different deductive coding sources for IAPT PWPs and patients. For interviews with PWPs we used the ‘theoretical domains framework’ (TDF) to help identify implementation barriers and faciliators^19^. For interviews with IAPT patients we used the ‘capability, opportunity, and motivation - behaviour change model’ (COM-B model) that is designed for characterising and designing behaviour change interventions^20^. Inductive codes were data-driven, and remained close to participants language where possible.

Two researchers read each transcript (GT and KS), and listened to the audio recordings before coding the transcripts. The data were coded in four phases: 1) We started with inductive line-by-line coding. After coding three transcripts, two researchers involved in coding (GT and KS) compared labels and agreed a set of codes to apply to all subsequent transcripts. 2) Codes were grouped into categories providing a working analytical framework. 3) We then deductively coded concepts from the COM-B and TDF models to the data where appropriate; some data were coded both inductively and deductively (See appendix). 4) Overarching themes and subthemes were developed based on what was necessary for intervention development.

N-Vivo software was used to apply the working analytical framework for phases 1 and 2, in which the framework was applied by indexing subsequent transcripts using the existing categories and codes. For phases 3 and 4 we used Microsoft Word and Excel.

### Patient and public involvement

During study conceptualisation, the research aims and design were reviewed by the UK Centre for Tobacco and Alcohol Studies Smokers’ Panel for feedback. In general, the study’s concept was well received, understood, and thought to be an important area of research. We consulted with the UK Centre for Tobacco and Alcohol Studies Smokers Panel and the Elizabeth Blackwell Institute’s Patient and Public Involvement Panel to develop the interview schedules, asking for guidance on topics and question phrasing.

### Research team and reflexivity

Personal characteristics: GT conducted the interviews, coding and analysis, and is a behavioural scientist. KS coded and analysed the interviews, and is a research assistant with expertise in health psychology. AS had oversight of coding and analysis, and is a qualitative methodologist.

Relationship with participants: A working relationship was established with IAPT PWPs and stop smoking advisors prior to the interviews. There was no relationship with IAPT patients prior to the interviews.

## RESULTS

### Participant profile

We interviewed 11 PWPs, six IAPT patients, and six smoking cessation advisors (Appendix for participant characteristics) from IAPT and smoking cessation services in the Midlands and South West regions of the UK. All twelve PWPs who were invited to participate, agreed. Eight IAPT patients were invited to participate, one did not reply, and one declined. All six smoking cessation advisors who were invited agreed to participate.

PWPs worked across two large NHS trusts in England, and ranged from newly qualified to senior PWPs. IAPT patients were seeking treatment for social phobia and mixed anxiety and depression, and were all regular daily smokers. Stop smoking advisors worked in a county council supported and privately-led smoking cessation services.

Below we present five themes, and 10 subthemes, with illustrative quotes.

### Theme 1: People use smoking to cope with mental health difficulties

Theme 1 characterises patients’ motivations to smoke, some of these motivations were automatic, and some were reflective; these motivations align deductively with the COM-B model^20^.

#### Smoking as a coping strategy

IAPT patients reported psychological benefits of smoking, including using smoking as a crutch in the face of trauma, as a form of stress relief, and as a comfort during difficult times. There was evidence that smoking to cope was an automatic motive for smoking, a “habit” or a “trigger”.

> “*I did a good nine months using* (smoking cessation medicine) *but then something quite traumatic happened … I just literally used smoking as a crutch really to kind of deal with it. It* (smoking) *was just kind of like an old habit that I fell back into because I felt like smoking had got me through other situations in life*.*”*
>
> − Female IAPT patient, aged 44-years, mixed anxiety and depressive disorder
>
> *“*(I smoke) *particularly if there’s quite a short deadline at work or something or if I get into an argument with somebody like the missus or something like that – an extreme argument not just the normal ones and then that might trigger it* (smoking).*”*
>
> − Male IAPT patient, aged 32-years, generalised anxiety disorder

#### Smoking as a form of self-harm

Smoking was described as a form of self-harm, either to replace “destructive” behaviours like drinking alcohol, or as existing on a continuum from unhealthy lifestyle behaviours (i.e., under-exercising, overeating, smoking), to self-injury. Smoking as a form of self-harm was described after reflecting on motivations for smoking.

> *“…it* (smoking) *might be kind of like a weird form of like self-harm in a way as well ‘cause when I … initially started smoking for maybe like a month whether I was drinking or not and I realised that that was also kind of like a coping mechanism but it’s very self-destructive and I realised there was no way that I was gonna be able to carry on doing that and stay in university and then I guess I kind of started smoking when I was going out. I was like ‘Okay well I’ll just stop drinking because that’s not something I’m gonna be able to like maintain’, but like smoking is a more socially acceptable way of having that kind of like destructive coping mechanism…”*
>
> − Female IAPT patient, aged 26-years, social phobia

One participant described that she uses self-harm to “take the edge off”, and that self-harm can replace smoking.

> *“I was just sad all the time, I just wanted to have a cigarette because I thought there was nothing in life for me. I have done self-harming to myself as well … which does take the edge off so if I’m self-harming myself I won’t have a cigarette*.*”*
>
> − Female IAPT patient, aged 60-years, mixed anxiety and depressive disorder

PWPs compared tobacco use and self-harm, describing self-harm as a continuum.

> *“… if we were to think about … kind of self-harm and self-injury, and there’s quite a big scale of that, so self-harm starting with maybe, um, under-exercising or overeating or smoking, and that then going up to kind of self-injury. So, I suppose, when we think about if people are smoking, are they smoking to cope in some way with difficult emotions or what’s going on? Um, so is it as easy as we just start smoking, or is there more going on there that would need to be replaced or worked on in some way, um, I think, maybe. Um, and is it just a bit of a break from life, you know, what… what is it they’re… obviously, there’s addiction, but is there something else, a bit more, that’s… um, needs to be thought about, in a way…”*
>
> − Female PWP, aged 27-years, 1-year experience as a PWP

### Theme 2: Smoking as a vicious cycle

Theme 2 “Smoking as a vicious cycle” was inductively developed as IAPT works predominately using a cognitive behavioural treatment (CBT) model for depression and anxiety but maps to the behaviour change wheel and COM-B model.

#### How people experience the cycle

IAPT patients and PWPs described how tobacco addiction related to thoughts, feelings, behaviours, and physical sensations, and discussed how these four domains interrelated. This subtheme is relevant for using a CBT cycle as a psychoeducational tool, and training PWPs on how tobacco fits into a CBT cycle worsening mental health.

One IAPT patient explained that smoking produced a feeling of relief, diminishing the physical craving, which led to feeling “calmer” but recognised that this may be nicotine withdrawal.

> *“If I have one* (cigarette) *it’s sort of a bit of a relief, not only just maybe five, ten minutes to myself to think it’s more that my body is getting what it’s craving as well and it just eases off and it’s a calmer feeling anyway but that’s probably me just not recognising the fact that I don’t get the nicotine or something”*
>
> − Male IAPT patient, aged 32-years, generalised anxiety disorder

One IAPT patient described smoking as a cycle of feeling relief and calm, followed by physical craving.

> *“Well it just makes me feel less anxious. Makes me feel a little bit calmer but then obviously you know you’re craving the next cigarette; you know you’re craving the next hit to keep you feeling calm*.*”*
>
> − Female IAPT patient, aged 44-years, mixed anxiety and depressive disorder

#### How PWPs perceive the cycle

During interviews PWPs automatically started to think about smoking in terms of a CBT cycle and how thoughts, feelings, behaviours, and physical sensations are interlinked. This theme was deductively developed based on the knowledge and skills domains of the TDF (‘theoretical domains framework’)^19^.

> *“If they’re already having physical problems they’re already internally focussing on their breathing, or palpitations…then smoking it can then add to the* (anxiety) *symptoms… because it* (smoking) *increases your palpitations and mimics the symptoms of anxiety… It’s definitely what we classify as an unhelpful behaviour*.*”*
>
> − Female PWP, aged 28-years, 1-years’ experience as a PWP
>
> *“I do think that smoking can be used as an um safety behaviour particularly in anxiety so people with tend to use that as a way to reduce their anxiety… they’re artificially bringing their anxiety down which is unhelpful. So, I guess in some ways understanding the habit of smoking and getting them to reduce that in the situation with anxiety could be helpful as well. Also, I think in terms of fitting it in and around with interventions that work with depression* (like behavioural activation) *I think that would probably work quite well*.*”*
>
> − Female PWP, aged 28-years, 4-years’ experience as a PWP

#### IAPT patient ‘buy-in’

near the latter end of the interviews the interviewer (GT) presented research showing that stopping smoking is associated with mental health benefits as large as taking anti-depressants, and asked what people’s thoughts were about this finding. All IAPT patients responded positively to this message. This theme maps on to the use of “education” as an intervention technique to influence motivation to quit by challenging automatic beliefs and promoting reflection^20^.

One person mentioned that framing smoking cessation messaging in this way might motivate her to try and quit.

> *“Stopping smoking as a treatment for mental health would make me more likely to want to do it* (quit smoking). *I think if it was framed more as like quitting smoking can actually help your mental health, not like they just wanna like shoe horn it in ‘cause that’s the kind of idea that I kind of had initially that it was like ‘Okay this is the population that we know smoke a lot, we want people to stop smoking so let’s kind of like shoe horn the stop smoking in with the therapy to try and cut down the general population*.*’”*
>
> − Female IAPT patient, aged 26-years, social phobia

Another person reflected on possible underlying mechanisms in the association between quitting smoking and improved mental health.

> *“I mean quitting smoking is gonna make your physical health better, which I guess it would probably make your mental health better”*
>
> − Female IAPT patient, aged 44-years, mixed anxiety and depressive disorder

When further prompting, and explaining that researchers believe that mental health improves upon quitting smoking because of breaking the tobacco addiction cycle, many IAPT patients reflected on their own mental health and tobacco addiction in the context of this hypothesis.

> *“Maybe the withdrawal symptoms are causing my anxiety sometimes*.*”*
>
> *“Yeah so when I’m clear of it* (tobacco addiction), *like clear of wanting one, out of the habit of going to one and it’s like been a lot of weeks maybe a month or two down the line, I do notice, yeah I’m a lot calmer and a lot happier actually. I have noticed that in the past when I’ve given up for a considerable period*.*”*
>
> − Male IAPT patient, aged 32-years, generalised anxiety disorder

### Theme 3: IAPT as a natural infrastructure for offering smoking cessation treatment

The Theoretical Domains Framework (TDF) was used to develop Theme 3^19^. We identified barriers and both positive and negative organisational culture towards smoking cessation treatment for people with depression and anxiety in current stop smoking services. PWPs were confident in their capability to offer smoking cessation treatment to IAPT patients.

#### Therapeutic pessimism and stigmatising attitudes towards helping people with mental health difficulties to quit

Interviews with stop smoking advisors were conducted to learn about behavioural techniques that they use to support people with common mental health difficulties to quit smoking. We instead found therapeutic pessimism and identified stigmatised views about helping people with mental health difficulties to quit smoking tobacco.

For example, one smoking cessation advisor inadvertently suggests people with severe depression are somehow reprehensible and cannot therefore stop smoking.

> *“Let’s say somebody has depression serious severe depression. They’re not going out, they’re not eating, their whole life has stopped, and they’re smoking. They just like to sit and smoke, and showing severe depression, so I feel that bringing them back to life will take a good amount of effort, however the effort to quit smoking may be too much effort in their life”*
>
> − Female smoking cessation advisor, aged 28-years, 2-years’ experience

Smoking cessation advisors reflected on the difficulty of treating people with common mental difficulties because they are “less self-motivated” and struggle with commitment.

> *“More often than not we sign these people* (people with common mental difficulties) *up anyway, even if they chronically relapse and may come back into the service. The support we provide behaviourally is based around motivational interviewing and trying to get them to provide the motivation. Whereas in these patients, the motivation doesn’t come from within…it doesn’t last… they’re less self-motivated, and so as a result it’s much harder to carry them on the course*.*”*
>
> − Male smoking cessation advisor, aged 35-years, 4-years’ experience
>
> *“Their* (people with common mental difficulties) *biggest problem is commitment. And a lot of things that actually become into depression and anxiety actually starts at the beginning, when they start to fail with commitments: things like, ‘Don’t worry, I’ll do that tomorrow’; but that never happens*.*”*
>
> − Male smoking cessation advisor, aged 39-years, 2-years’ experience

Another smoking cessation advisor mentioned that smoking helps people with mental illness “stop thinking about their problems”.

> *“They smoke ‘cause of enjoyment. It’s to stop them from thinking about their problems. That’s why they smoke more than people who don’t have mental illness. That’s why they’re really, really difficult to work with*.
>
> − Female smoking cessation advisor, aged 34-years, 2-years’ experience

#### PWPs express their ability to provide behavioural support

During interviews PWPs referred to a behavioural change models and techniques, and described how they use these to motivate people to make lifestyle changes when patients had low motivation. They described low motivation as the “voice of depression”.

> *“Somebody I spoke to the other day said, ‘well I, I did quit but since I’ve started feeling more stressed again, I’ve started smoking again’. So (s/he) might think um, ‘I’m failing at this - I’ve started smoking again’, and that might feed into more negative cognitions they might have about themselves… and equally one of the main things um about depression is just that sense of demotivation, and getting people to recognise that’s the voice of depression and that’s what maintains the depression*.*”*
>
> − Female PWP, aged 27-years, 2-years’ experience

PWPs described using models like COM-B to assess patients’ desire to change behaviour; once the patients’ capability, opportunity and motivation is determined, the PWP uses this to inform treatment planning.

> *“If they wanted to change it* (smoking)… *there has to be a desire to change it, so in the assessment we’re checking three factors: their capability to change, the opportunity they’ve got to change and the motivation they’ve got to change their behaviour. So, if they’re keen to change it, then we would look at integrating it and look at making those changes. If they’re not in a place where they can change, then we look at kind of trying to motivate them to get to that point*.*”*
>
> − Female PWP, aged 30-years, 4-years’ experience

During interviews, PWPs described using motivational interviewing techniques to explore pros and cons of behaviours to encourage change and re-engagement when motivation is low.

> *“Um, so with obviously motivation um, we try to sort remind people of the pros and cons of change so that okay um, what do you – what’s wrong with your current situation. Do you want to make a change and what’s that change worth and er sometimes that can make people re-engage*.*”*
>
> − Female PWP, aged 32-years, 3-years’ experience
>
> *“I think it would be harder because of their low motivation and I think that’s why exploring the beliefs around it might be helpful and um exploring with them their readiness for change, so in terms of their motivation, when somebody is thinking about making a change, they go through different stages, a pre-contemplation, a contemplation and then a stage where they’re ready to change, and if they’re ready to change it* (smoking cessation intervention) *would be really good because you move them from pre-contemplation to action*.*”*
>
> − Female PWP, aged 28-years, 4-years’ experience

Another PWP explained that they use behavioural activation to help people with low mood make behavioural changes.

> *“if they have low mood, one of the techniques we look at is called behavioural activation, and we look at managing or changing their behaviours and increasing the positive behaviours to get more of a balance… So it depends what they want to work on, what their goals are for treatment, but it can be worked on and integrated into the therapy if necessary, because if their lifestyle factors are impacting their mood and they aren’t eating properly, or exercising or they are kind of drinking a lot to manage, (we) have to adapt that behaviour in order to move forward, so it definitely can be integrated in*.
>
> − Female PWP, aged 30-years, 4-years’ experience

#### Integrating smoking cessation support into IAPT treatment

All PWPs thought that integrating smoking cessation support into IAPT would complement IAPT’s organisational culture/climate, and that it could be logically integrated into the current treatment model.

One PWP highlighted that smoking cessation treatment would “sit really nicely in the IAPT service”.

> *“I think it’s a really good idea because we work on sleeping. We work on eating. We work on exercise. We work on caffeine. We work on all the elements of somebody’s wellbeing and the only thing we don’t really touch is smoking, which is – we even work on alcohol use so to have treatment with us, you have to be below the alcohol limits. So actually, we look at a lot of these different things to treat people. Smoking seems to be the only one we don’t really touch, so that’s – I think it would sit really nicely in the IAPT service*.*”*
>
> − Female PWP, aged 28-years, 1-years’ experience

One PWP linked smoking to other “safety behaviours” that she helped IAPT patients manage.

> *“So, um, it, it’s often I suppose similar to many other kinds of safety behaviours. We term safety behaviours, or responses that people might have as something that feels better in the short term, but in the long term can make you feel worse physically but also it can fuel more negative cognitions*.*”*
>
> − Female PWP, aged 27-years, 2-years’ experience

Another PWP reflected on integrating smoking cessation into usual care, and highlighted that one option would be to focus on smoking during IAPT treatment planning, identifying a treatment goal, so that smoking cessation support would be integral to the patient’s treatment plan.

> *“Thinking about how that applies I guess to an intervention around smoking, it’s about identifying that as a goal in your first session. So, we have a treatment planning session, where we plan treatment with them collaboratively, so I think using things like the hot cross bun to be able to identify if smoking is a coping behaviour, or if smoking is related in any way, or if smoking is just something that they do, um, and then using that to sensitively question well is it something you want to look at or a goal that you want to think about*.*”*
>
> − Female PWP, aged 28-years, 4-years’ experience

The interviewer noted that someone who stops smoking will usually experience tobacco withdrawal symptoms like low mood and anxiety, and asked how this could fit into a service that aims to help improve people’s mental health. PWPs accepted this challenge and thought that the experience of tobacco withdrawal symptoms fitted well as the IAPT model does ask patients to engage with “evidence-based” treatments that can make people “feel worse before they feel better”.

> *“As long as* (patients) *are aware that they will potentially feel worse and that they will have those withdrawals and it will be difficult but we’re here to support them. The key part is making them aware and then it’s their choice what they do. I don’t think there’s a problem with us doing that to someone or encouraging someone to* (quit smoking) *when they’re in mental health support. Because… the end goal is still to get them to feel better in the long-term… and if they can do that whilst they’re in our support I think that’s probably better…”*
>
> − Female PWP, aged 30-years, 4-years’ experience
>
> *“We are telling people to make changes and sometimes those changes can be quite significant and, quite distressing for the patient… sometimes people do feel worse before they feel better. So, we always tell people at the beginning of the treatment, that this is not going to be easy. That this is um evidence-based, it has good outcomes the treatments that we offer but… you might feel worse in the beginning before the treatments start to take effect*.*”*
>
> − Female PWP, aged 32-years, 3-years’ experience

### Theme 4: Risk management

Theme 4 aligns with multiple TDF domains: knowledge, skills, social/professional role and identity, beliefs about capabilities, and environmental context and resources.

All PWPs raised the concept of “risk” during Interviews, and described that assessing and managing patient “risk” was a routine part of their role, and they were confident in their skills and ability to assess risk. PWPs quickly applied their professional role in managing risk to psychological withdrawal symptoms from tobacco. PWPs indicated that “risk management” was especially relevant to smoking cessation treatment, as if a patient feels that they need smoking to cope, or if they fail to quit smoking and feel worse, the patient will be supported if their mental health deteriorates.

> “*if* (patients) *are feeling worse* (mentally), *then it’s a safe place for them to tell us. We assess their risk every single session, we ask if they have any thoughts of suicide, any action taken, any plans to end their life; we look at thoughts of self-harm, any action taken, that kind of stuff, so essentially check how risky they feel. Um, thoughts are normal and we kind of normalise that to the patient. We say, ‘You know, if you’re standing on the train platform, um, you might have thoughts of “What if I jumped in front of that train?” Doesn’t mean you’re gonna act on it. You know, nine out of ten people have had that thought. I’ve had that thought*.*”*
>
> − Female PWP, aged 30-years, 4-years’ experience
>
> *“…we check in each appointment… because we know that things can change. Either if somebody is, somebody’s mood is worse. Somebody’s anxiety increases, um and that’s something that we might need to revisit. I’ve worked with people before who might initially at assessment have seemed okay, but when you get through for their appointment, or over the course of therapy… they feel worse, and maybe they start self-harming again, or having thoughts of suicide… because you’ve got a chance to revisit* (risk) *each time, then you might need to change treatment direction. You might say we need to send you to the adult mental health team… or maybe we need to think about high intensity therapy”*
>
> − Female PWP, aged 27-years, 2-years’ experience

### Theme 5: Intervention refinement and evaluation

#### PWP training requirements

All PWPs agreed that they had the skills, and had perceived confidence in their capabilities. PWPs identified that they needed training on the “evidence-base” around smoking and mental health.

> *“I think we’ve got the skill base, it’s just that sort of research and evidence base*.*”*
>
> − Female PWP, aged 32-years, 3-years’ experience

She continued to identify specific topics for training.

> *“I guess the effects of the different medications, patches and so on and maybe the differences between e-cigarettes… more information about those things so that we…know that we’re giving people facts. We’re an evidence-based service…”*
>
> *“*(I would like) *to know about physiology of um withdrawal. Are they more or less likely to get shortness of breath? Or do their airways as they get unblocked… do we encourage them to exercise at the same time, or we would focus on just smoking cessation and then start an exercise*.*”*
>
> − Female PWP, aged 32-years, 3-years’ experience

Another PWP identified the importance of having key messages in training.

> *“Statistics about relationships between depression and anxiety and smoking. The key studies. The key points to take home, to drive home to patients would be really important… to know the effect on different disorders*.*”*
>
> − Female PWP, aged 28-years, 1-years’ experience

#### Messages for commissioners

When exploring organisational barriers to implementing smoking cessation treatment in to IAPT PWPs identified some service and trust-level barriers that needed addressing

PWPs emphasised the importance of careful messaging to IAPT patients.

> *“I don’t want people to be put off accessing mental health services because they think that we’re gonna jump in on telling them to stop smoking and that sort of thing, um, which I don’t think it would be that anyway, but just being conscious of that, I suppose…”*
>
> − Female PWP, aged 27-years, 1-years’ experience

PWPs highlighted the impact of austerity on IAPT, and that integrating smoking cessation treatment might require a change in treatment session duration, or a reduction in case-load.

> *“One big problem that we have is sort um too many people in not the right step of the Step Care model because of cuts made in the psychological services and complex needs services, so – which means that we see a lot of complex patients which actually this kind of telephone guided self-help is not necessarily suitable for so then we might need to have longer session or more sessions or treat them for longer or um, it might get more complex so um, that’s a major um barrier*.”
>
> − Female PWP, aged 32-years, 3-years’ experience
>
> *“Um, maybe if, I’m not fully sure, the only one would be maybe if we’re offering longer appointments or having treatment for a bit longer, it maybe that we aren’t then seeing as many people as we would before because we’re holding onto those people for a bit longer so that could maybe implement the service in terms of numbers of people accessing treatment*.*”*
>
> − Female PWP, aged 30-years, 1-years’ experience

## DISCUSSION

People with common mental illness used tobacco smoking as a coping strategy, and reported that smoking had several psychological benefits, including as a ‘crutch’ in the face of trauma, as a form of stress relief, and as a comfort during difficult times. Other qualitative and quantitative studies have identified these patterns in similar populations^21,22^. Our study provided new insights into the idea that tobacco is used as a form of self-harm, as described by people with common mental illness and by psychological wellbeing practitioners (PWPs). A common theme identified throughout the interviews was that tobacco addiction was described as a vicious cycle. This study provides an understanding into how naturally people with common mental illness can see how tobacco addiction is related to their thoughts, feelings, behaviours, and physical sensations, and how PWPs automatically integrated the tobacco withdrawal cycle into this CBT model, without prompting from an interviewer. IAPT (a psychological service) PWPs and patients recognised how the tobacco withdrawal cycle mimics common mental illness symptoms, how tobacco withdrawal may worsen mental health symptoms, and how breaking this cycle could benefit mental health.

An important finding of this research is that PWPs appear to be very different from other mental health professions in terms of their understanding about how smoking and mental health are related^10,14,15^. PWPs appear to have positive attitudes to implementing smoking cessation treatment into psychological services for people with common mental illness. For example, Sheals and colleagues conducted a mixed methods systematic review and meta-analysis of mental health professionals’ attitudes to treating smoking cessation in people with mental illness. The most commonly held beliefs were that patients were not interested in quitting, and that quitting smoking is too much for patients to take on. This review included studies of professionals working predominately in services that implemented medically-based treatment models^10^.

During the interviews it emerged that IAPT may be a more appropriate place to offer smoking cessation than local ‘stop smoking’ services where people with common mental illness would usually be referred.^23^ We found that the smoking cessation advisors we interviewed had stigmatised views about helping people with mental health difficulties to quit smoking. This is curious as their training involves specific modules about helping this population to quit, and breaking down common myths about smoking cessation in this population^24^. We cannot ascertain that these views are generalisable to other smoking cessation services, however a survey of smoking cessation advisors found similar attitudes^25^. This finding does indicate a potential barrier for people with depression/anxiety when attempting to quit smoking via local ‘stop smoking’ services; and signifies the importance of PWPs as experts in applying evidence-based behavioural methods to help people with depression/anxiety to make lifestyle changes.

PWPs all had an in-depth understanding that mental and physical health were inter-related, thus naturally they accepted the idea of offering an integrated smoking cessation treatment into IAPT for patients who were interested in receiving help to quit. PWPs accepted the notion that stopping smoking may induce psychological withdrawal symptoms and did not see this as a problem to psychological treatment in IAPT. PWPs noted that often in IAPT patients “get worse before they get better”, and that they deal with this concept often. PWPs regularly described how risk is routinely assessed and managed in IAPT.

PWPs made suggestions for service leads and commissioners. It was noted that if smoking cessation treatment was to be offered in IAPT, service leads should carefully frame this as patients may be deterred from mental health treatment if smoking cessation treatment was perceived as mandatory. PWPs also reflected on the impact of austerity on IAPT services. PWPs noted that if smoking cessation treatment were to be integrated into IAPT that longer treatment sessions, or smaller case-loads might be necessary. Given that smoking cessation interventions are among the most cost-effective health care interventions available^26^, and that smoking cessation is linked to considerable mental health benefits^11^, offering smoking cessation as part of routine mental health care seems sensible from a patient- and commissioning-perspective.

In this study we used snowballing strategy for recruitment because PWP and smoking cessation advisors have a particularly high caseload, and recruitment via recommendation from service managers overcame this. However, this recruitment method may introduce bias into our sample as service mangers potentially invited only motivated and receptive team members. It should also be noted that there was some variability in the depth of interviews conducted with IAPT patients; telephone interviews tended to be less reflective and more descriptive.

### Clinical implications

An important finding from these interviews was that IAPT patients and psychological wellbeing practitioners (PWPs) welcomed the idea of smoking cessation as a treatment for mental health. They were able to reflect on their own experiences and identify examples of when tobacco withdrawal has mimicked anxiety or depression symptoms, and how this relates to their mental health difficulties. PWPs naturally were able to link tobacco withdrawal to a CBT vicious cycle in which thoughts, feelings, behaviours, and physical sensations are interrelated. PWPs clearly identified the clinical implications of using the cycle as a psychoeducational tool during intervention sessions.

Based on these interviews, it might be possible to integrate smoking cessation treatment into psychological services like IAPT, using a variation of NCSCT’s standard treatment programme for smoking cessation^24^, whereby PWPs focus on addressing the relationships between smoking, the withdrawal cycle and links to mental health, using a CBT model (Example here: https://www.youtube.com/watch?v=HiYBGOQ-PIo&t=8s).

An unexpected finding was that stop smoking advisors had pessimistic and stigmatising attitudes towards helping people with common mental health difficulties to quit. It is important that stop smoking services complete the “smoking and mental health” module provided by NCSCT^27^.

## CONCLUSION

PWPs have positive attitudes towards smoking cessation treatment for people with common mental illness. IAPT PWPs and patients accept new evidence that smoking tobacco may harm mental health, and quitting might benefit mental health. PWPs have expertise in helping people with depression/anxiety to make lifestyle changes in the face of stress, anxiety, low mood, and poor motivation. IAPT appears to be a natural environment for smoking cessation intervention; however, there may be service level barriers.

**Table 1.** Summary of themes and subthemes.

| Theme | Subtheme | Description |
| --- | --- | --- |
| People use smoking to cope with mental health difficulties | Smoking as a coping strategy | People who smoke describe how they use smoking to cope with stress and common mental health symptoms. |
|  | Smoking as a form of self-harm | People who smoke describe how they use smoking to self-harm. |
| Smoking as a vicious cycle | How people experience the cycle | People who smoke discuss how tobacco addiction fits into a vicious cycle that is linked to their mental health. |
|  | How PWP's perceive the cycle | Examples of how PWP's interpret smoking as a vicious cycle |
|  | IAPT patient 'buy-in' | The interviewer presents evidence that smoking may improve mental health, equal to taking anti-depressants – people who smoke respond to this evidence and how this makes them feel about their addiction. |
| IAPT as a natural infrastructure for offering smoking cessation treatment | Therapeutic pessimism and stigmatizing attitudes towards helping people with mental health difficulties to quit | Views of smoking cessation practitioners about helping people with common mental illness quit smoking. |
|  | PWP's express their ability provide behavioural support | PWP's give examples of their skills and how they motivate people with common mental illness to make behavioral changes to improve their overall well-being. |
|  | Integrating smoking cessation support into IAPT treatment | Examples of how treating tobacco addiction fits in well with the current IAPT ethos, i.e. physical and mental health are interlinked, patient-centered approach, transparency. |
| Risk management |  | PWP's explain how one of their main roles is to assess and manage patient risk like suicidal ideation. |
| Intervention refinement and evaluation | PWP training requirements | PWP's state what their training needs are to deliver an integrated smoking cessation treatment in IAPT. |
|  | Messages for commissioners | PWP's reflect on top-level facilitators and barriers to delivering an integrated smoking cessation treatment into IAPT. |

## Data Availability

Anonymised transcript data will be uploaded to the University of Bath's Research Depository, and will be restricted access data, made available only through the data guardian. These details will be released soon.

